# BRIEF COMMUNICATION: High level of Anti SARS-Co-V2 RBD Antibody one year post booster vaccine hospital workers in Indonesia; Was second booster needed?

**DOI:** 10.1101/2023.02.15.23285958

**Authors:** Amila Hanifan Muslimah, Marita Restie Tiara, Hofiya Djauhari, Hafizh Dewantara, Evan Susandi, Agnes Rengga Indrati, Bachti Alisjahbana, Arto Yuwono Soeroto, Rudi Wisaksana

## Abstract

Healthcare workers in Indonesia acquired a complete 2 doses of Sinovac in early 2021 and first booster dose of Moderna in July 2021. In August 2022, the ministry of health prioritized healthcare workers to acquire the second booster dose of Moderna as antibody levels from the year before may have waned. We conducted a sequential serosurvey aimed to determine the level of SARS CoV2 S-RBD antibody reached by the first vaccine, after the first booster, and before the second booster to understand the dynamics of the antibody level. COVID-19 antibody test was conducted using the FastBioRBD^tm^ test with a maximum limit detection level of 4000 BAU/mL. First serosurvey which was conducted in June 2021, one to six months after Sinovac vaccination, showed a median antibody level of 41.4 BAU/mL (IQR 10 – 629.4 BAU/mL). The second serosurvey was conducted one month (August 2021) after the first Moderna booster vaccine, and showed a median level of 4000 BAU/mL (IQR 3081 – 4000 BAU/mL). While the last serosurvey conducted a year (August 2022) after the booster, showed 4000 BAU/mL (IQR 4000 – 4000 BAU/mL). Only 39 (11.9%) healthcare workers have antibody levels below the maximum level of 4000 BAU./mL We did not see the waning of antibody levels among healthcare workers approximately 1 year after the booster. It increases perhaps due to the natural infection caused by the omicron variant outbreak in early 2022. Based on this fact, we suggest considering if the second booster dose is really necessary. The limited vaccine supply can better be given to the person or other high-risk groups of patients who has a low level of antibody based on serological testing.

## Introduction

SARS-CoV-2 infection has become a health problem worldwide, including in Indonesia. (1) Presence of anti-SARS-CoV-2 antibody post-infection or due to vaccination has been confirmed to provide some protection against infection and even more against severe COVID-19 disease. (2) Vaccination is therefore mandatory. However, current evidence is still lacking about how many boosters are necessary for healthcare workers and the community at large. It is even more difficult to determine because anti-SARS-CoV2 antibody level does affect by the type of vaccine we received, exposure to natural infection as well as time since last vaccination/exposure, and internal factors like age and comorbidity. (3,4)

Healthcare workers were greatly affected during the first wave in 2020 and the Delta strain of SARS CoV-2 outbreak in July-September 2021. Our healthcare workers acquired complete 2 doses of Sinovac vaccine from January to June 2021. Sinovac was chosen as the vaccine used for healthcare workers because it was most readily available in Indonesia at that time. Next, they acquire Moderna Booster in August 2021. During the Omicron outbreak in early 2022, many health care workers were also infected with COVID-19 but no severe infection was rarely observed. In July 2022, the Indonesian Ministry of Health decided to prioritize a second mRNA vaccine for healthcare workers in Indonesia. (5,6)

With the vaccine scheme that we acquired and the periodic exposures to COVID-19 during outbreaks, we came to wonder how high is our anti RBD-Level? And, is this second booster really necessary? Information about the level of Anti SARSCoV2-SRBD antibody can assist in evaluating the need for additional vaccination. (7) This study aims to describe the SARS CoV2 S-RBD antibody level among Hospital Workers in three consecutive serosurveys at different time points which can provide information on whether a booster is necessary.

## Materials and methods

This research was conducted at Hasan Sadikin General Hospital as the top referral hospital for COVID-19 in West Java, Indonesia, with high exposure of COVID-19 infection healthcare workers during the pandemic. This study received approval from the Health Research Ethics Committee of Hasan Sadikin General Hospital Number LB.02.01/X.6.5/117/2022. It is a single-center, observational analysis study with a prospective design using primary data examining levels of SARS CoV 2 IgG S RBD antibody in hospital healthcare workers. Data consisted of basic participant information from interviews and results of antibody levels for SARS-COV-2 IgG S-RBD. All participants were vaccinated using Sinovac in the months January-June 2021.

Our first serosurvey was conducted in July 2021, right before the third vaccine dose or the first booster using the Moderna vaccine. The second serosurvey was conducted in August 2021, one month after the provision of the first Moderna booster Vaccine. The third serosurvey was conducted in August 2022, 12 months after the first Moderna booster, right before the second Moderna booster that was programmed by the Ministry of Health.

Eligible participants were health workers of Hasan Sadikin General hospital collected by voluntary participation. All participants were vaccinated with the complete two doses of the inactivated virus vaccine (Sinovac) and one booster dose of mRNA-1273 (Moderna) full dose. This study used a convenience sampling method. All hospital staff who followed these vaccination schemes and were willing to participate in the study were included in the survey.

We use the following operational definition. Physicians were all MDs, mostly specialists but also included general practitioner working in the hospital. COVID-19 wards were areas that had the highest risk of spreading COVID-19, including COVID-19 intensive care, general care, and emergency isolation room. History of close contact with confirmed cases were face-to-face contact with probable cases or confirmed cases within one-meter radius and 15 minutes or more, direct physical touch with probable or confirmed cases, or people who provided direct care to probable or confirmed cases without using standard personal protective equipment. Last COVID-19 infections (having previous COVID-19 infections in between surveys) were a person who has positive RT-PCR results or rapid antigen resulta with at least three symptoms of COVID-19 infection based on the interview.

Determination of anti-SARS-CoV2 S-RBD Antibody uses a point of care quantitative immunochromatographic assay (FastBioRBD^tm^) produced by Wondfo, Guangzhou Biotech, China which was rebranded for distribution by PT Biofarma, Indonesia (Persero). This equipment has a value range between 1 to 200 Arbitrary Unit (AU) where 1 is regarded as the minimal level for being seropositive. We later multiplied the result by 20 to obtain the standard WHO binding antibody unit (BAU). Therefore, 20 BAU/mL was the bottom limit for seropositivity, while the maximum detection limit was 4000 BAU/mL. A similar fluorescent immunoassay test (Wondfo Finecare anti-RBD) has been utilized and tested in Qatar. (8). To validate the FastBioRBD^tm^ test, 71 randomly selected samples were tested using the GenScript cPass SARS-CoV-2 Neutralization Antibody Detection Kit (Genscript Biotech, Leiden, Netherlands). Briefly, serum samples as well as negative and positive controls were diluted 1:10 in sample dilution buffer, mixed 1:1 with HRP-RBD working solution, and incubated at 37 °C for 30 min. For more information sees in reference (9).

The baseline characteristics of participants described were factors that might influence SARS CoV2 S-RBD antibody level, which was age, gender, occupation, comorbidity, last COVID-19 infection, work zone, history of close contact, and levels of SARS-CoV-2 Ig G S-RBD. Data on baseline characteristic were obtained based on information from the interview with a validated questionnaire. Anti-SARS CoV2 S-RBD antibody level was described in the descriptive table as well in graphical form.

## Results

On the first survey, we enrolled 570 person during the first booster vaccination day. After blood collection, these subjects directly acquired the first booster of Moderna (full dose). One month after this first booster we enrolled 355 person in the second survey. One year after the first booster, we enrolled and collect serum samples from 330 persons in the third survey right before the second booster of Moderna (figure). Participants’ demography and characteristics are summarized in the Table.

**Table.**
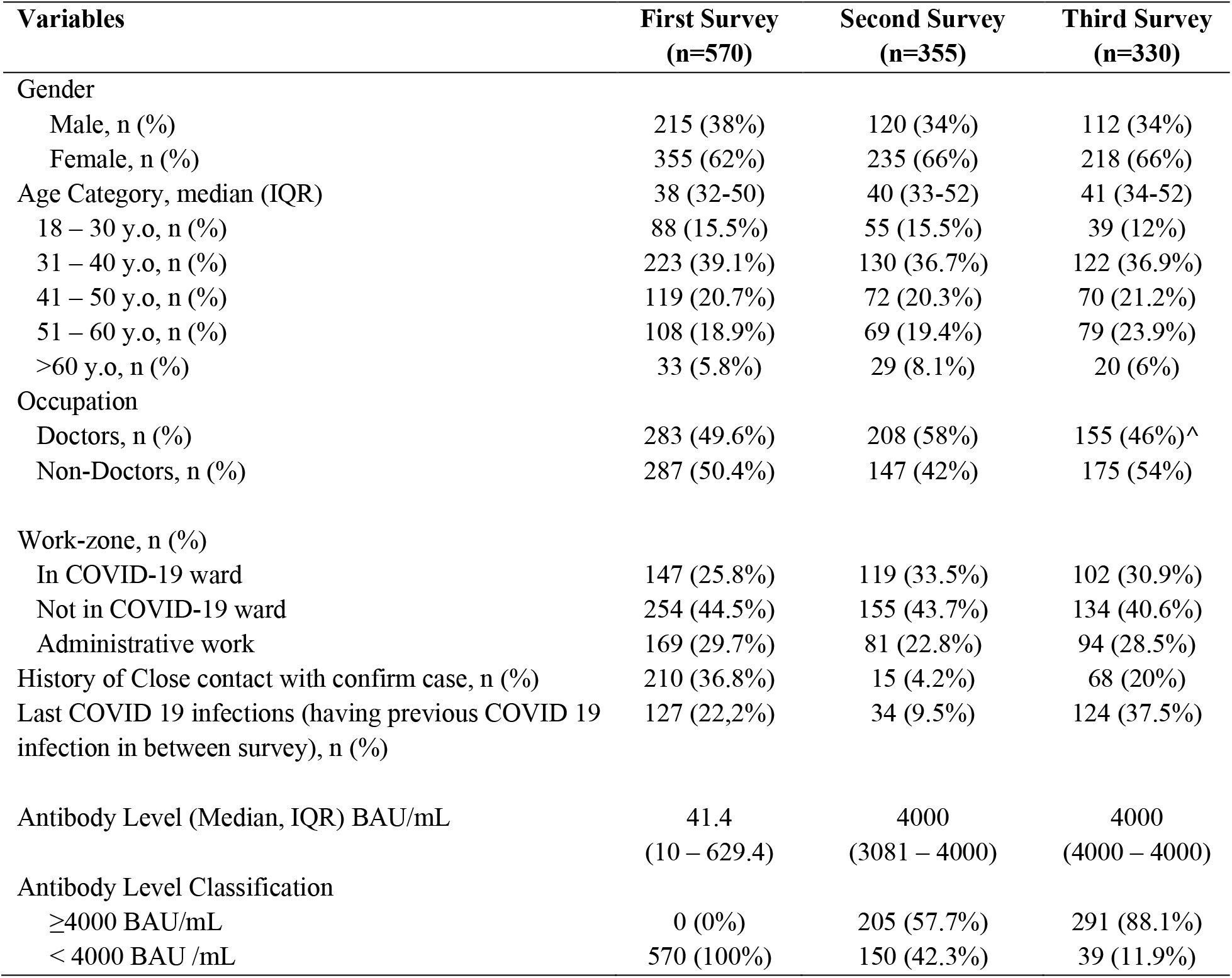
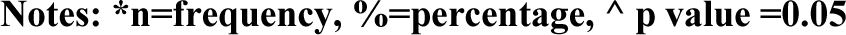
Baseline Characteristics of Participants.

**Table. Baseline Characteristics of Participants**
| Variables | First Survey<br>(n=570) | Second Survey<br>(n=355) | Third Survey<br>(n=330) |
| --- | --- | --- | --- |
| Gender |  |  |  |
| Male, n (%) | 215 (38%) | 120 (34%) | 112 (34%) |
| Female, n (%) | 355 (62%) | 235 (66%) | 218 (66%) |
| Age Category, median (IQR) | 38 (32-50) | 40 (33-52) | 41 (34-52) |
| 18 – 30 y.o, n (%) | 88 (15.5%) | 55 (15.5%) | 39 (12%) |
| 31 – 40 y.o, n (%) | 223 (39.1%) | 130 (36.7%) | 122 (36.9%) |
| 41 – 50 y.o, n (%) | 119 (20.7%) | 72 (20.3%) | 70 (21.2%) |
| 51 – 60 y.o, n (%) | 108 (18.9%) | 69 (19.4%) | 79 (23.9%) |
| >60 y.o, n (%) | 33 (5.8%) | 29 (8.1%) | 20 (6%) |
| Occupation |  |  |  |
| Doctors, n (%) | 283 (49.6%) | 208 (58%) | 155 (46%)^ |
| Non-Doctors, n (%) | 287 (50.4%) | 147 (42%) | 175 (54%) |
| Work-zone, n (%) |  |  |  |
| In COVID-19 ward | 147 (25.8%) | 119 (33.5%) | 102 (30.9%) |
| Not in COVID-19 ward | 254 (44.5%) | 155 (43.7%) | 134 (40.6%) |
| Administrative work | 169 (29.7%) | 81 (22.8%) | 94 (28.5%) |
| History of Close contact with confirm case, n (%) | 210 (36.8%) | 15 (4.2%) | 68 (20%) |
| Last COVID 19 infections (having previous COVID 19 infection in between survey), n (%) | 127 (22,2%) | 34 (9.5%) | 124 (37.5%) |
| Antibody Level (Median, IQR) BAU/mL | 41.4<br>(10 – 629.4) | 4000<br>(3081 – 4000) | 4000<br>(4000 – 4000) |
| Antibody Level Classification |  |  |  |
| ≥4000 BAU/mL | 0 (0%) | 205 (57.7%) | 291 (88.1%) |
| < 4000 BAU /mL | 570 (100%) | 150 (42.3%) | 39 (11.9%) |

Notes: \*n=frequency, %=percentage, ^ p value =0.05

**Fig 1.**
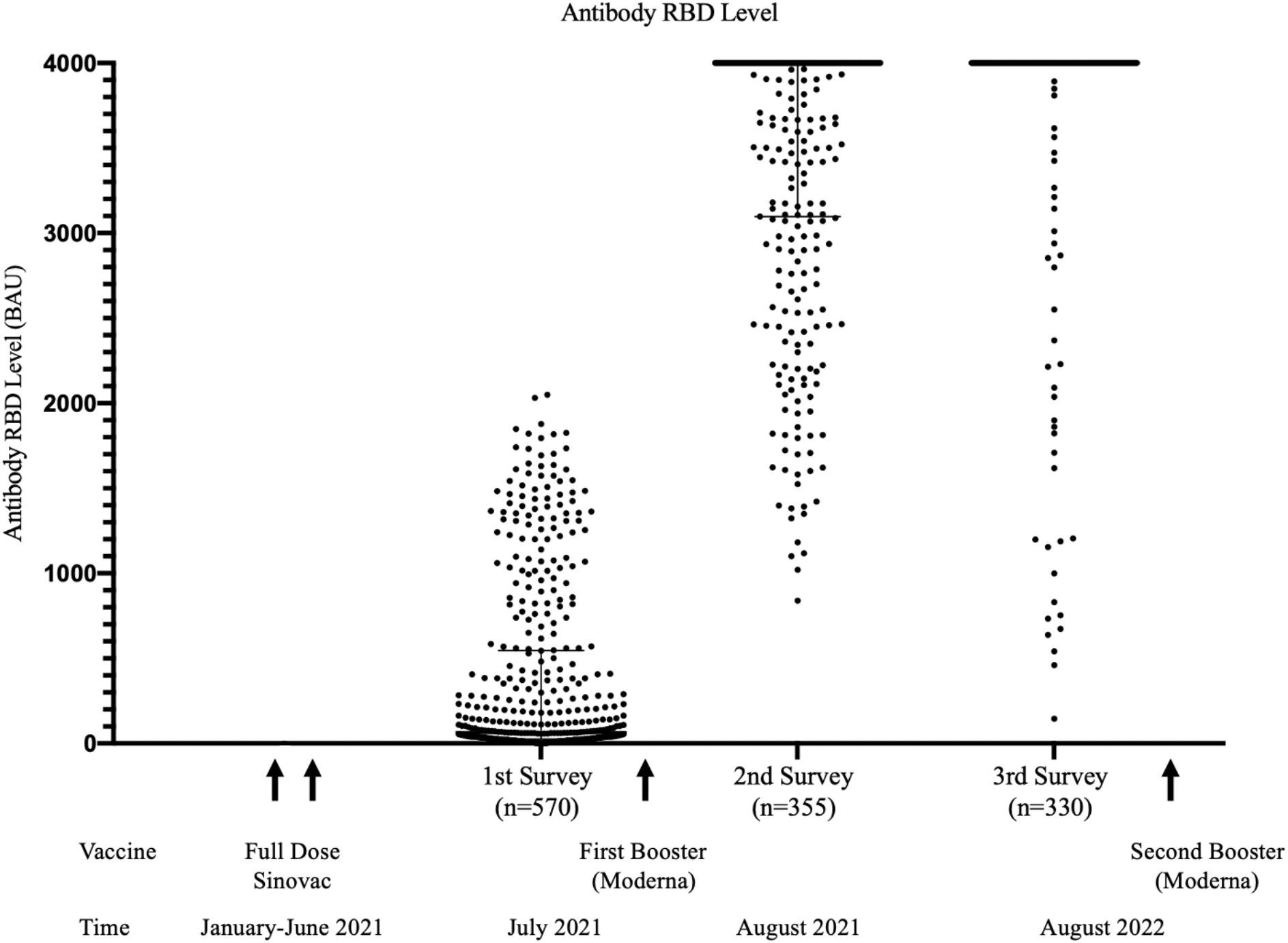
SARS CoV 2 S RBD Antibody Level Across Serosurveys. Sequential serosurvey of SARS CoV2 S-RBD antibody before first booster Moderna, one month after first booster Moderna, and 12 months after first booster Moderna. Most healthcare workers have very high level of SARS CoV2 S-RBD antibody level 12 months post third COVID 19 vaccine dose. Second Moderna booster was given right after the blood taking off the third survey.

The median age of the participants was 41 (IQR 34 to 52) years old, and 46% were practicing physicians. 64 (19.3 %) were specialists, and 91 (27.5%) were general physicians. Among the non-physician, 55 (16.6%) were nurses, 10 (3.03 %) were laboratory staff and 73 (22.1%) were the administrative staff. Most of the participants belong to the non-covid ward group (254, 44.5%), while 147 (25.8%) were stationed and routinely care for COVID-19 patients in the ward.

During the first survey, the SARS CoV2 S-RBD antibody level median was 41,4 BAU/mL (Range 0.2 – 2.049.6 BAU/mL). One hundred eight (32.7 %) participants showed negative results for SARS-CoV-2 S-RBD antibody (BAU/mL < 20) and 175 (30.7%) participants did not reach the median value (< 41.4 BAU/mL). No participant reached a maximum detection level of 4000 BAU/mL. From the interview, 127 (%) reported having confirmed COVID-19 illness since the beginning of the Pandemic.

In the second survey, we observed a median of 4000 BAU/mL (Range 840,4 – 4000). One hundred fifty (42.3%) participants had antibody levels below the maximum detection level of 4000 BAU/mL. No participant showed negative anti-SARS CoV2-SRBD. Within one month duration between the first and second survey, 34 participants reported having confirmed covid illness.

In the third survey, we obtained a median SARS CoV2 S-RBD antibody level of 4000 BAU/mL (range 144,8 – 4000 BAU/mL). In this survey 124 (37.5%) participants reported having confirmed COVID-19 illness within one year between the second and the third survey. The distribution of antibody levels based on the time of examination can be seen in the Figure.

Individuals with maximum antibody levels >4000 BAU/mL one month after 3^rd^ vaccination had more COVID-19 infections after 3^rd^ vaccination, with 57 participants (58.2%) and antibody levels >4000 BAU/mL twelve months after 3^rd^ vaccination having more COVID-19 infections with 106 participants (85.5%). The less infection in the second survey probably was due to the shorter time interval between the first survey and the second survey.

A bivariate analysis test was performed on factors potentially related to antibody levels on the last serosurvey. Bivariate analysis showed that being a physician was associated with a lower SARS CoV2 S-RBD antibody levels. Doctors have lower antibody levels (4000, Range 460-4000 BAU/mL) compared to non-doctors professions (4000, Range 145-4000 BAU/mL) (p =0.05). The multivariate analysis gave the same result, which was that being a doctor was a factor that had a significant result (adjusted OR 0.512 (95% CI 0.258-1.015) S1Table. Antibody levels twelve months after the third vaccination did not reflect a protective effect against COVID infection 19.

Seventy-one randomly selected serum samples underwent validation tests with Genscript C-Pass. FastBioRBD^tm^ compared to C-Pass showed a correlation with r-value of 0.78 (95% CI 0.678-0.862) S2 Figure.

## Discussion

We found an increasingly high antibody levels among healthcare workers in our hospital one year post booster. We think that this was due to natural exposure throughout the pandemic. Many believe that the antibody level that has been stimulated by the COVID-19 vaccine may gradually decline within several months (10,11). However, our observation did not show this situation.

Antibody levels were examined in participants about six months after being vaccinated with two Sinovac vaccinations shown to reach a marginal level. There is still a substantial proportion of participants who showed negative serology. A study in Turkey conducted in March 2021 observing Anti-RBD antibody levels 28 days post two doses of the Sinovac vaccine in 330 health workers showed that 99.4% of health workers had a seropositive result with a median of 154.78 BAU/mL. While only thirty-two-point-four percent of participants have maximum antibody levels. (12) Antibody examination on healthcare workers in Hasan Sadikin General Hospital has a median time of 5,5 months after the second dose of Sinovac vaccination, thus showing lower results. This is similar to a study conducted by Fonseca et al in Brazil, in September 2021, that antibody levels 6 months after two doses of the Sinovac vaccine lower levels with a median of 66.7 BAU/mL and 16 (2%) participants became seronegative. (13) The difference of these findings can also be affected by the local epidemic dynamics in each country/region. (14)

The increase in antibody levels one month after receiving the first booster of Moderna vaccination was expected. Over half of the respondents showed a maximum readable level of RBD antibody in this survey. However, there were still a few subjects who showed only a moderate increase in antibody response. Adding a booster of mRNA vaccine in subjects who have acquired inactivated vaccine was shown to increase their antibody level. A study in Chile has shown that the group of subjects who acquired two Sinovac vaccines and a booster with the BNT162b2 RNA vaccine had higher antibody levels 1 month after the third vaccination compared to the group given two Sinovac vaccines and the third vaccine with Sinovac. (15) Several studies have also shown the effect of the Moderna vaccine, which demonstrates a higher antibody response than the BNT vaccine.(16,17)

The third survey was conducted in August 2022, one year after the last booster of the Moderna vaccine and after we went through the Omicron outbreak in early 2022. In this survey, we found an even higher anti-RBD antibody level among hospital staff and there were no signs of weaning of antibody level. The increased antibody level between the second survey to the third survey was most probably due to COVID-19 exposure during the Omicron outbreak. The result of our study is similar to the national serosurvey conducted in Indonesia. The Indonesian serosurvey has been conducted routinely in October and November 2021 and July 2022. The serosurvey also reveals an increasing antibody level in all communities in Indonesia. The vaccinated person has a higher antibody response than a non-vaccinated person. (18,19) A similar study in Italy conducted in January-November 2021 also showed similar results where anti-COVID RBD antibody increases months after receiving a booster vaccine. (20)

Our study showed that participants with high antibody levels experienced more COVID-19 infection. As we know now from various reports, antibody level does not effectively prevent infection, especially with the new variant. (21,22) But it significantly lowers the chances of having severe covid-19 illness. (23) The higher infection among these groups could be due to their working environment or lifestyle which is already related to higher exposure to COVID-19 since the beginning, therefore they have higher antibody response, and chances of new infection. Our data and several other reports have consistently shown that (in developing countries) infections are mostly due to casual contact rather than from the working environment. (24) Whereas casual contact was less affected by the hospital protocol as well as national regulation.

Our study extensively uses the quantitative immunochromatographic assay which is easy to use and affordable. The test uses a cassette-based dry reagent with a small portable reader which can be utilized in a point-of-care setting. Compared to other tests, this test has good and acceptable validity. Our validation test of 71 samples with Genescript C-Pass showed a good correlation results. The Subject who has maximum detection level has a more than 90% result based on genscript SVNT assay had a sensitivity of 63.8%, specificity of 100%, positive predictive valueof 100%, and negative predictive value of 41.4%. A validation test in Qatar has selected 150 subject serum samples with r-value of 0.7 (95% CI 0.60 to 0.80). (8)

Our study has some limitation. We collect the person invited to the survey based on the person who was willing to be tested. Thus, the number of samples, especially on the second and the third survey may not represent the whole hospital staff population. However, within our observation, we have represented the whole range of types of workers and exposure levels. Information about the previous COVID-19 was collected through interview or self-reporting. This method may not be the most accurate as there might be information bias. Although based on other reports, self-reporting may lead to lower prevalence than registered reports. (18,19)

## Conclusion

We did not find a waning of humoral immunity to COVID-19 among healthcare workers in our hospital. Most probably this is due to the high transmission of SARS CoV2 and the outbreak of the omicron in early 2022. Most healthcare workers have a very high level of SARS CoV2 S-RBD antibody level 12 months post third COVID-19 vaccine dose. Based on this result, we should consider if a fourth vaccine dose is necessary for all. To make efficient use of the vaccine distributed, we suggest to incorporating antibody testing and only providing booster to those who have lower anti-RBD Antibody level.

## Data Availability

All relevant data are within the manuscript and its Supporting Information files.

## ACKNOWLEDGEMENT

We are grateful for the support from Wondfo, Guanzhou Biotech, China, and PT Biofarma Indonesia (Persero) for providing the FastBioRBD^tm^ fluorescent immunoassay reader and the rapid test reagents for our study. Thank you to Research and Innovation Indonesian Institute (BRIN) for providing initial operational support for conducting the study. We thank the director of Hasan Sadikin General Hospital for allowing this study to be conducted and the staff of the hosptital who participated in this study. We thank the Dean of the Medical Faculty of Padjadjaran University who a provide laboratory facilities for sample management and testing in this study.

## DATA AVAILABILITY

The data used to support the findings of this study were included in the article.

## Supplementary

**S1 Table. Multivariate Analysis of Antibody Levels 12 Months after The Third Vaccination**

**S2 Figure. Surrogate Virus Neutralization Test (SVNT) Correlation with Anti-RBD FastBioRBD**^**tm**^ **in Participants after 12 Months of Third Vaccination**

## REFERENCES

1. World Health Organization. Novel Coronavirus (COVID-19) Situation. 2022 Feb 25 [cited 25 February 2022]. Available from: https://covid19.who.int/.

2. Sadarangani M, Marchant A, Kollmann TR. Immunological mechanisms of vaccine-induced protection against COVID-19 in humans. Nat Rev Immunol. 2021;475–484.

3. Shrotri M, Navaratnam AMD, Nguyen V, Byrne T, Geismar C Fragaszy, et al. Spike-antibody waning after second dose of BNT162b2 or ChAdOx1 Lancet. 2021;398(10298):385–387.

4. Zeng G, Wu Q, Pan H, Li M, Yang J, Wang L, et al. Immunogenicity and safety of a third dose of CoronaVac, and immune persistence of a two-dose schedule, in healthy adults: interim results from two single-centre, double-blind, randomised, placebo-controlled phase 2 clinical trials. Lancet Infect Dis. 2021;S1473-3099(21):00681–2.

5. Minister of Health. Decree of the Minister of Health of the Republic of Indonesia. Technical Instructions for the Implementation of Vaccination in the Context of Combating the 2019 Corona Virus Disease (Covid-19) Pandemic; 2022 [cited 20 February 2022]. Database: Management of Public Information on the Supreme Audit Board [internet].Available from: https://peraturan.bpk.go.id/Home/Details/169665/permenkes-no-10-tahun-2021.

6. Ministry of Health of the Republic of Indonesia. Circular Letter Number Hk.02.02/C/3615/2022 Concerning the 2nd Booster Dose of Covid-19 Vaccination for Health Human Resources. Ministry of Health of the Republic of Indonesia. 2022 July 28 [cited 30 July 2022]. Available from: https://www.kemkes.go.id/downloads/resources/download/lain/Surat-Edaran-Vaksinasi-COVID_19-Dosis-Booster-2.pdf.

7. Lo Sasso B, Giglio RV, Vidali M, Scazzone C, Bivona G, Gambino CM, Ciaccio AM, Agnello L, Ciaccio M. Evaluation of Anti-SARS-Cov-2 S-RBD IgG Antibodies after COVID-19 mRNA BNT162b2 Vaccine. Diagnostics (Basel). 2021;11(7):1135.c

8. Shurrab FM, Younes N, Al-Sadeq DW, Liu N, Qotba H, Laith J. Abu-Raddad LJ, Nasrallah GK. Performance evaluation of novel fluorescent-based lateral flow immunoassay (LFIA) for rapid detection and quantification of total anti-SARS-CoV-2 S-RBD binding antibodies in infected individu. Int. J. Infect. Dis. 2022; 118:132–137

9. GenScript. cPass SARS-CoV-2 Neutralization Antibody Detection Kit Instruction for Use. In: Nanjing GenScript Diagnostics Technology Co. L, editor. 2022.

10. Shrotri M, Navaratnam AMD, Nguyen V, Byrne T, Geismar C Fragaszy, et al. Spike-antibody waning after second dose of BNT162b2 or ChAdOx1 Lancet. 2021;398(10298):385–387.

11. Zeng G, Wu Q, Pan H, Li M, Yang J, Wang L, et al. Immunogenicity and safety of a third dose of CoronaVac, and immune persistence of a two-dose schedule, in healthy adults: interim results from two single-centre, double-blind, randomized, placebo-controlled phase 2 clinical trials. Lancet Infect Dis. 2021;S1473-3099(21):00681–2.

12. Dinc HO, Saltoglu N, Can G, Balkan II, Budak B, Ozbey D, Caglar B, Karaali R, Mete B, Tuyji Tok Y, Ersoy Y, Ahmet Kuskucu M, Midilli K, Ergin S, Kocazeybek BS. Inactive SARS-CoV-2 vaccine generates high antibody responses in healthcare workers with and without prior infection. Vaccine. 2022;40(1):52–58.

13. Fonseca MHG, de Souza TFG, de Carvalho Araújo FM, de Andrade LOM. Dynamics of antibody response to CoronaVac vaccine. J Med Virol. 2022;94(5):2139–2148.

14. Zimmermann P., Curtis N. Factors That Influence the Immune Response to Vaccination. Cli Microbiol Rev. 2019;32(2):e00084–18.

15. Vargas, L., Valdivieso, N., Tempio, F. et al. Serological study of CoronaVac vaccine and booster doses in Chile: immunogenicity and persistence of anti-SARS-CoV-2 spike antibodies. BMC Med. 2022; 20:216.

16. Steensels D, Pierlet N, Penders J, Mesotten D, Heylen L. Comparison of SARS-CoV-2 antibody response following vaccination with BNT162b2 and mRNA-1273. JAMA. 2021;326(15):1533–1535.

17. Collier AY, Yu J, McMahan K, et al. Differential kinetics of immune responses elicited by Covid-19 vaccines. N Engl J Med. 2021;385(21):2010–2012.

18. Indonesian Health Ministry. National Survey Serology Results. 2022 March 18 [cited 24 January 2023] In: COVID 19 Indonesia [Internet]: Jakarta: Indonesian Health Ministry. Available from : https://covid19.go.id/artikel/2022/03/18/hasil-survei-serologi-covid-19-di-indonesia-november-desember-2021

19. Indonesian Health Ministry. Third National Serology Survey. 2022 September [cited 24 January 2023] In: Indonesian Health Ministry [Internet]: Jakarta: Indonesian Health Ministry. Available from : https://www.youtube.com/watch?v=cgESZbe9rC0.

20. Lo Sasso B, Agnello L, Giglio RV, et al. Longitudinal analysis of anti-SARS-CoV-2 S-RBD IgG antibodies before and after the third dose of the BNT162b2 vaccine. Sci Rep. 2022;12:8679.

21. Chakraborty C, Sharma AR, Bhattacharya M, Lee SS. A Detailed Overview of Immune Escape, Antibody Escape, Partial Vaccine Escape of SARS-CoV-2 and Their Emerging Variants with Escape Mutations. Front Immunol. 2022;9(13): 801522.

22. Cromer D et al. Neutralizing antibody titers as predictors of protection against SARS-CoV-2 variants and the impact of boosting: a meta-analysis. Lancet Microbe 2022;3:e52–61.

23. Agrawal U et al. Severe COVID-19 outcomes after full vaccination of primary schedule and initial boosters: pooled analysis of national prospective cohort studies of 30 million individuals in England, Northern Ireland, Scotland, and Wales. Lancet 2022;400: 1305–20.

24. Yu HJ, Hu YF, Liu XX, Yao XQ, Wang QF, Liu LP, Yang D, Li DJ, Wang PG, He QQ. Household infection: The predominant risk factor for close contacts of patients with COVID-19. Travel Med Infect Dis. 2020; 36:101809.

